# Experience with Social Distancing Early in the COVID-19 Pandemic in the United States: Implications for Public Health Messaging

**DOI:** 10.1101/2020.04.08.20057067

**Authors:** Ryan C. Moore, Angela Lee, Jeffrey T. Hancock, Meghan Halley, Eleni Linos

**Affiliations:** Department of Communication, Stanford University, CA, USA; Department of Dermatology, Stanford University, CA, USA; Department of Epidemiology, Stanford University, CA, USA

**Keywords:** coronavirus, COVID-19, natural language processing, public health messaging, social distancing compliance

## Abstract

Our goal is to inform ongoing public health policy on the design and communication of COVID-19 social distancing measures to maximize compliance. We assessed the US public’s early experience with the COVID-19 crisis during the period when shelter-in-place orders were widely implemented to understand non-compliance with those orders, sentiment about the crisis, and to compare across age categories associated with different levels of risk. We posted our survey on Twitter, Facebook, and NextDoor on March 14^th^ to March 23^rd^ that included 21 questions including demographics, impact on daily life, actions taken, and difficulties faced.^1^ We analyzed the free-text responses to the impact question using LIWC, a computational natural language processing tool^2^, and performed a thematic content analysis of the reasons people gave for non-compliance with social distancing orders. Stanford University’s IRB approved the study.

In 9 days, we collected a total of 20,734 responses. 6,573 individuals provided a response (≥30 words) to the question, “Tell us how the coronavirus crisis is impacting your life.” Our data (Figure 1) show that younger people (18-31) are more emotionally negative, self-centered, and less concerned with family, while middle-aged people are group-oriented (32-44) and focused on family (32-64) (all *p* values < .05 corrected for multiple comparisons). Unsurprisingly, the oldest and most at-risk group (65+) are more focused on biological terms (e.g., health-related topics), but were surprisingly low in anxiety and high in emotionally positive terms relative to those at lower risk.

We also content-analyzed 7,355 responses (*kappa’s* > .75) to the question, “What are the reasons you are not self-isolating more?” Of these participants, 39.8% reported not being compliant, with the youngest group (18-31) having the lowest compliance rate (52.4%) compared to the other age groups (all > 60%; all *p* values < .01). Table 1 describes the seven primary themes for non-compliance. Non-essential work requirements, concerns about mental and physical health, and the belief that other precautions were sufficient were the most common reasons, although other rationales included wanting to continue everyday activities and beliefs that society is over-reacting. Childcare was an important concern for a subset of respondents.

Overall, our findings suggest that public health messages should focus on young people and 1) address their negative affect, 2) refocus their self-orientation by emphasizing the importance of individual behavior to group-level health outcomes, and 3) target the specific rationales that different people have regarding the pandemic to maximize compliance with social distancing.

---

**Figure 1.**
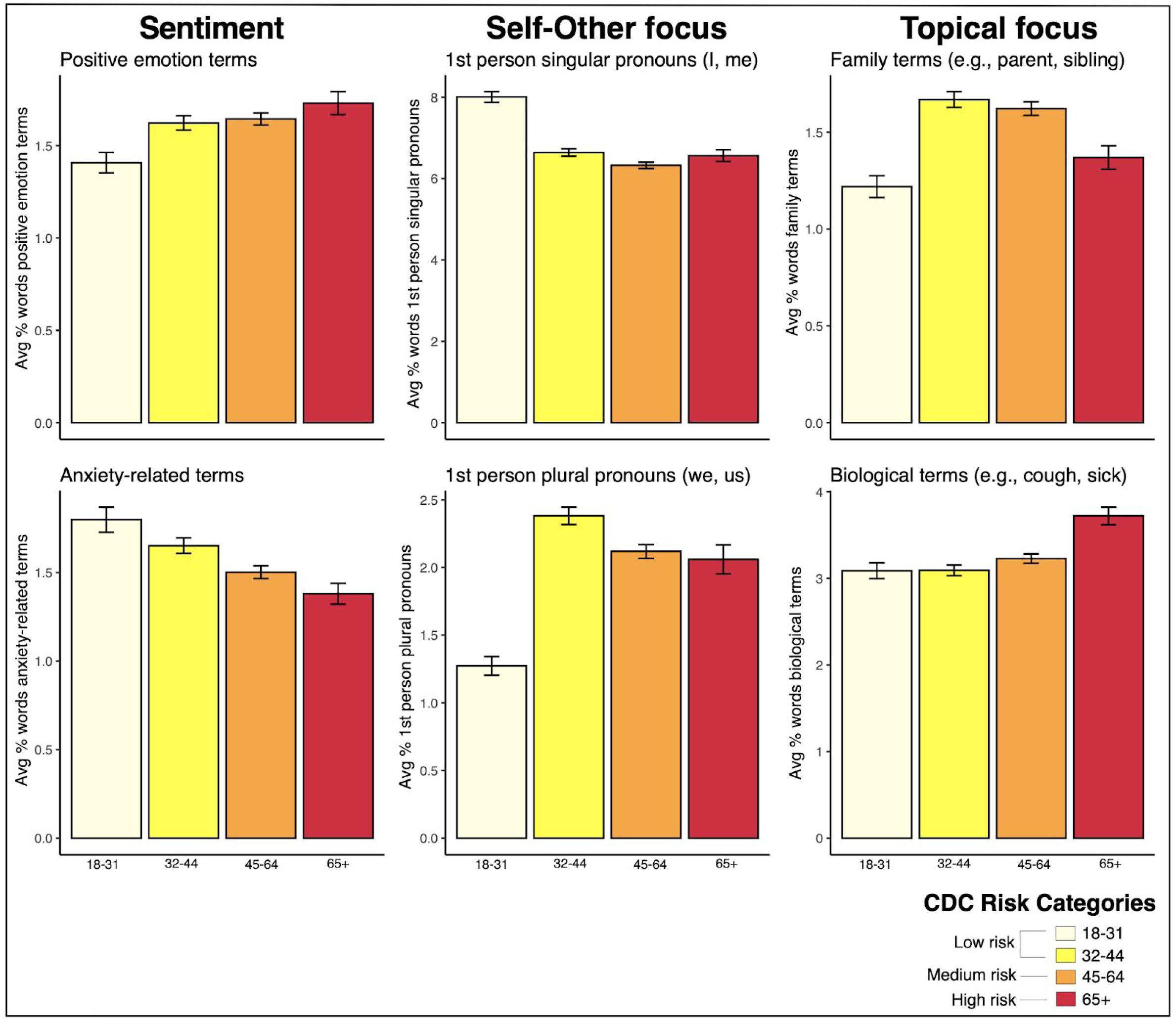
Mean number of words in language categories, by age group. Age groups are modeled after the age groups reported by the Center for Disease Control in their summary of COVID-19 cases in the US.^3^ Bars represent standard errors.

**Table 1.** Primary Themes for Non-Compliance and Messaging Strategies

| Theme | Example | % Freq | Messaging Strategy to Increase Compliance |
| --- | --- | --- | --- |
| Non-Essential Work | "Work is not canceled, if I don't go I'll lose my job." | 28.2 | Convey forthcoming government support; Communicate necessity of remote work solutions to employers |
| Mental & Physical Health | "Total self isolation would probably drive me to suicide." | 20.3 | Recognize hardships, identify safe well-being activities |
| Taking Sufficient Precautions | "I already wash my hands regularly and cover my mouth when I cough or sneeze. I am not concerned with catching [the] virus." | 18.8 | Explain why stronger measures required; Convey norms about compliance |
| Non-Essential Activities | "Some appointments are in-person. Need to see friends sometimes." | 13.9 | Provide clarity on what activities are essential |
| Society Over-Reacting | "I think the news media was making everyone panic and overreact." | 12.7 | Emphasize danger to all if society is compromised |
| Childcare-Related | "Really really hard to do with little kids - I'm reducing a lot of contact, but not all." | 4.8 | Construct parent-specific recommendations |
| Government-Related | "There has been a lack of clear direction on when one should start to seriously isolate." | 1.4 | Develop consistent messaging from federal, state and local levels of government |

## Data Availability

Data are available from the authors upon request.

## Acknowledgements

We would like to acknowledge the individuals who helped with the survey dissemination online.

## Footnotes

1 Nelson LM, Simard JF, Oluyomi A, et al. US Public Concerns About the COVID-19 Pandemic From Results of a Survey Given via Social Media. JAMA Intern Med. Published online April 07, 2020. doi:10.1001/jamainternmed.2020.1369

2 Pennebaker, J. W., Boyd, R. L., Jordan, K., & Blackburn, K. (2015). The development and psychometric properties of LIWC2015. (https://repositories.lib.utexas.edu/bitstream/handle/2152/31333/LIWC2015_LanguageManual.pdf)

3 Severe Outcomes Among Patients with Coronavirus Disease 2019 (COVID-19) — United States, February 12–March 16, 2020 (https://www.cdc.gov/mmwr/volumes/69/wr/mm6912e2.htm)

